# Using Artificial Intelligence to Personalize Caring Contact Messages for Recently Discharged Patients: Protocol for a Mixed-Methods Feasibility Study

**DOI:** 10.1101/2025.02.12.25322159

**Authors:** Rosalie Steinberg, Jasmine Amini, Abigail Wiliszewski, Prudence Po Ming Chan, Zardar Khan, Anne L. Martel, Kaleigh Starritt, Mark Sinyor, Ayal Schaffer

**Affiliations:** Department of Psychiatry, Sunnybrook Health Sciences Centre, Toronto, ON, Canada; Sunnybrook Research Institute, Toronto, ON, Canada; St. John’s Rehabilitation Centre, Toronto, ON, Canada; Institute of Medical Science, University of Toronto, Toronto ON, Canada

## Abstract

Suicide risk is substantially elevated following discharge from a psychiatric hospitalization. Caring Contact (CC) messages are brief messages of hope, support and information sent post-discharge that can improve mental health outcomes, including suicidal ideation and behaviours. Patients who received CCs in previous studies indicated a desire for increased personalization. To support personalization in an efficient and scalable manner, we will pilot the training of a Large Language Model (LLM) that constructs tailored CCs using information extracted from patients’ electronic health records (EHRs) created during the index psychiatric hospitalization. This is a three-phase, mixed-methods study. In phase 1, psychiatric inpatient clinical staff members will be recruited to generate personalized CC messages from representative, de-identified samples of EHRs. This will be considered the control CCs. Clinical staff focus groups will then identify key components of a successful CC message. In phase 2, the control CCs and focus group feedback will be used to train an LLM on a large number of EHRs to produce personalized CC messages. The LLM will be given the same EHRs used in phase 1 to produce CC messages. In phase 3, we will recruit 30 patients with lived experiences (PWLE) of psychiatric hospitalization to evaluate and compare the control CCs with the LLM-generated messages to determine the acceptability of AI-generated CC messages. We hypothesize that LLM-generated CC messages will achieve acceptability ratings at least as positive as the control CC messages. Since CC messages are modifiable and can be altered to suit the needs of various clinical settings, the findings of this study can potentially be broadly generalizable.

## Introduction

Suicide risk is substantially elevated during the first 90-days post-discharge from a psychiatric hospitalization,^1,2^ especially for those experiencing suicidal ideation.^1,3^ This may, in part, owe to the fact that individuals may experience worsening symptoms when they return to daily life stressors and are adjusting to the loss of or diminished contact with healthcare providers.^1^ One key element to establishing continuity of care is to support patients’ mental health outcomes following discharge.^4^ However, post-discharge interventions can be costly, and maintaining continuity during the transition period from inpatient to outpatient care may not be feasible for many institutions. Therefore, there is a need for easily adaptable and cost-effective interventions that can bridge gaps in care and decrease suicide risk following discharge from a psychiatric hospitalization.

### Caring Contact Messages

CC messages are brief communications of hope, support and resources sent to patients following discharge from hospital. Since first being studied in the early 1970s, CCs have been trialled globally, with several studies reporting positive effects of their use on patient mental health including reducing psychiatric readmissions,^5^ suicidal ideation,^6^ and suicidal behaviour.^6,7^ A recent meta-analysis on the effectiveness of CCs with respect to suicide-related outcomes produced mixed results.^8^ Although CCs were associated with a reduction in suicide attempts during the year following discharge, they did not consistently reduce suicide mortality rates, nor did they reduce hospitalizations for suicidal behaviour during the same period.^8^ The latter finding can be difficult to interpret since requiring hospitalization can be a positive sign of accessing needed care and help-seeking, but also a negative sign of increased necessity for an acute care admission. Nonetheless, these findings may, in part, reflect the significant heterogeneity between study methodologies and samples as well as the CC interventions themselves. CC messages are built to foster connectedness, which can strengthen perceptions of social support, and act as a protective factor against suicidal thoughts and behaviours, among other adverse outcomes.^8^ This emphasizes one of the mediating effects of CCs, which is to encourage connectedness and adherence to mental health care, as well as help-seeking behaviours among high-risk populations.

### A Previous RCT and QI Project for Caring Contact Messages

Our research group piloted sending CC messages in a recent randomized control trial (RCT) comparing mental health outcomes between those who received and did not receive a series of CCs following discharge from a major teaching hospital in Toronto, Canada.^9^ Our results indicated that CCs attenuated the worsening of mental health symptoms observed in the group who received the CC compared to the control group. Results from this RCT were then used to inform a patient-centred Quality Improvement (QI) project, which used focus groups to incorporate patient and community member feedback to enhance the acceptability of our initial CC before piloting these adapted messages amongst a sample of recently discharged patients. Although underpowered to compare mental health symptoms pre- and post-discharge, participants reported a mean drop in depressive symptoms with the updated CC messages.^10^ A majority of our sample also expressed that CCs helped them feel more hopeful about their recovery and encouraged them to seek further help if needed.

The main finding from this recent QI project indicated patient preference for increased personalization of CC messages.^10^ Participants consistently identified a desire to have messages addressed directly to them from members of their clinical care team, and to incorporate specific information from their inpatient stay.^10^ Examples of relevant content could include gains made over the course of their admission, specific reminders of positivity in their life (e.g., family, pets), and reminders of their individualized safety plan (developed with their clinical staff) during moments of crisis.^10^ While providing personalized care, especially within mental health interventions, is beneficial, creating individualized content can be expensive and time-consuming.

### Utilizing Artificial Intelligence to Enhance Caring Contact Messages

Artificial Intelligence (AI) is commonly described as the making of intelligent machines; since intelligence is usually considered a human trait, the term “artificial” denotes that a machine or computer has such a trait.^21^ Reviews of the usage of AI in digital mental health interventions have identified personalization of content as a notable strength when considering the efficacy of these interventions.^11,15^ The use of AI in mental health care is burgeoning, particularly as a component of mobile mental health interventions. A recent scoping review found AI-based mental health interventions such as the use of mobile health apps to be feasible, generally well received, and effective at reducing symptoms of poor mental health (e.g. mood, anxiety symptoms).^11^ However, there was significant heterogeneity between studies and the majority of included samples were small.

Although the usage of AI in mobile health treatments/chatbots is ever-expanding, little research has explored its use in one-way patient messaging (from provider to patient) within a psychiatric context. An example of one-way AI-generated patient messaging could be accomplished by Large-Language Models (LLM). LLM is a type of AI that offers a unique opportunity to produce personalized CC messages, thus fulfilling patient needs while remaining cost-effective and efficient. LLM uses machine learning to understand and generate empathetic human language and is typically trained on large amounts of data.^13^ Preliminary work in other fields, such as primary care and diagnostic imaging, indicates that AI-based messaging systems can promote adherence to follow-up care and are capable of producing clear, relevant draft messages for patients.^12,13^ Further work and consultation with care providers is needed to refine these programs for day-to-day use.

As AI and its applications in healthcare continue to evolve, there are significant moral, ethical, and legal considerations regarding its use that cannot be overlooked. Foremost, AI poses considerable privacy concerns. To develop personalized messages to patients, an AI program would necessarily require access to patient EHRs and sensitive personal health information (PHI). This is a particular concern for institutions that collaborate with external commercial providers who may retain PHI and/or collate this data across jurisdictions to produce large datasets.^14^ There are also notable concerns regarding the use of AI chatbots or messaging for mental health care as these programs can deliver misinformation, nonsensical information (sometimes termed AI “hallucinations”), or harmful information.^15^ Whether AI can be incorporated into mental health supports while maintaining the ethical and legal standards of our healthcare system remains an open question.

In summary, AI, and specifically LLMs, offer an opportunity to enhance CC messaging and facilitate access to continuing support/resources in the high-risk post-discharge period. The current study will explore the appropriateness of AI-generated CC messages by training a LLM, compared to control messages written by clinical staff members. A cautious approach that assesses the feasibility and acceptability of using AI to produce CC messages is a necessary step to begin to determine whether AI technology can be harnessed to improve patient health while also keeping patient safety at the forefront.

### Objectives

The following are the objectives of the study:

1. To explore the feasibility of training an LLM by incorporating feedback into the extraction template and training model, to create personalized CC messages.
2. To examine the acceptability of and attitudes towards AI-generated CC messages as compared to control staff-generated CC messages among those with lived experience of a psychiatric hospitalization.

## Method

### Study design

This is a multi-phase iterative project. In phase 1, we will recruit clinical staff members from the inpatient psychiatric ward to create control CC messages. The messages will be reviewed by study personnel to ensure relevant and appropriate information from patient EHRs are included. To prepare for the creation of CC messages by the LLM, study personnel will also identify types of information from patient records which should be included in the messages.

This will guide the LLM on which information needs to be selected from a patient’s electronic health record to create CC messages. In phase 2, our AI specialists (ZK, AM) will train the LLM on a sample of approximately 100-200 patient EHRs. In phase 3, community members/patients with lived experience of psychiatric hospitalization (PWLE) will evaluate the tone, style, and efficacy the CC messages produced by AI compared to control messages.

#### Phase 1

##### Participants

A representative sample of 10 care team members from the inpatient psychiatric unit at Sunnybrook Health Sciences Centre, University of Toronto, will be recruited to participate in phase 1. The goal is to ensure the representation of participants from across the clinical care spectrum, including psychiatrists, social workers, occupational therapists, recreational therapists, nurses, learners, and members of the spiritual care team.

##### Inclusion Criteria

For inclusion in phase 1, participants must be a member of the clinical care team on the inpatient psychiatric unit at Sunnybrook Health Sciences Centre with experience in the care of psychiatric inpatients. All staff are age 18 or above, and able to read and understand English.

##### Procedure

Participation in phase 1 involves two stages: CC message creation by team member dyads followed by focus group discussions. All staff participants will be presented with a de-identified, sample patient EHR. With this “sample record,” each dyad will be asked to craft a personalized CC message from a representative sample of randomized and de-identified patient EHRs. To ensure a diverse range of patient records are identified for the creation of CC messages, a comprehensive array of variables will be used to randomize the selection. These criteria include: gender, adult age at admission, length of stay, and final diagnosis. We will also ensure that all patient records are from 2023 and include responses to the Columbia Suicide Severity Rating Scale. Study personnel will de-identify the patient EHRs that are randomly selected as samples. Before the clinical staff begin creating the CC messages, study personnel will develop guidelines to identify important information for inclusion in the messages. Basic structure and length constraints, as well as an emphasis on increased message personalization, positive and hopeful emotional resonance and empathy, will be given as guidelines. Participants, however, will have full control of message content.

Once staff participants have crafted their CC, two focus groups will be assembled consisting of recruited clinical staff members, a study co-primary investigator from psychiatry, our AI development leader, and a community representative, from our mental health partner peer-support agency, with both lived experience of mental illness and mental health research expertise. Using guidelines developed by various members of the study team, the focus group will discuss the components of a successfully personalized CC message from the perspective of the clinical staff members. Discussing the strengths and weaknesses of the CCs written by care staff members will be instrumental in developing the control CC messages, which will effectively include personalized message content.

**Figure 1.**
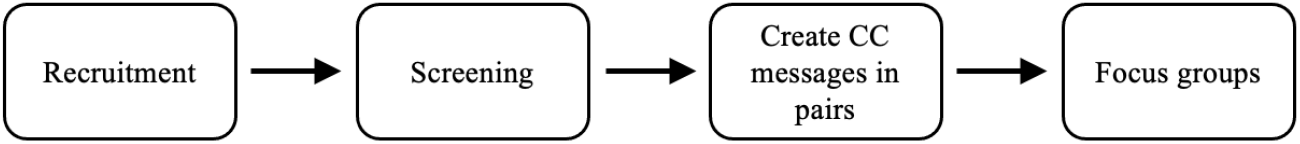
Phase 1 Procedures. *Note*. Procedures to be completed during phase 1 of the study.

#### Phase 2

##### Procedure

Data from the focus groups will be transcribed and thematically analyzed. Further, data from both stages of phase 1 will be used by our AI developers to craft an extraction template. Study personnel will also use a text labeller to manually identify elements from electronic patient health records to include in the LLM. The template will specify what information needs to be extracted from patient electronic health records by the LLM and will be used to train the model. We will input sections of the clinical notes into a Bidirectional Encoder from Transformers (BERT) model to extract the relevant details needed to populate a CC message template. BERT,^16^ developed by Google, is a language model that has shown excellent performance in multiple natural language processing tasks including named entity recognition, which classifies and extracts entities within text into predefined categories (e.g., reason for visit or employment status).^17^

Once the relevant information has been extracted and inserted into a CC message template, we will use Llama 2, a LLM released publicly by Meta,^18^ to edit this template to ensure a more natural message flow. Llama 2 generates text in response to a prompt (i.e. in instruction-tuned variation) and has been shown to perform well in a wide variety of tasks (e.g., dialogue use cases).^18^ The personalized messages from phase 1 will be used to develop prompts that adjust the “tone” of the message produced by the LLM so that it mimics the persona of a clinical care team member.

Our proposed pipeline will employ two types of language models to achieve quality CC generation. The first type is a representation model, such as BERT, which is specialized in understanding and classifying text rather than generation. The second type is a generative model, such as GPT or Llama, which excels in producing human-like text. This dual-model approach will allow us to identify important clinical information and generate a CC with a natural flow.

###### Stage 1 - Entity Identification

The initial stage will focus on determining which broad categories of clinical entities should be included in a CC to meet the standards defined by our focus group discussions. In this stage, 10 examples will suffice to help us identify the high-level categories that make up quality CC messages.

###### Stage 2 - Named Entity Recognition Implementation

Named Entity Recognition (NER) is a technique that allows for automatically identifying and classifying specific information within text. While stage 1 identifies broad categories, stage 2 will focus on detecting specific instances of these categories within text. This gradual recognition requires the development of a dataset to train a BERT model.^22^ For example, if “employment status” is an entity category identified in stage 1, the BERT model would learn to identify specific text snippets such as “currently employed at grocery store” or “on disability leave” as instances of employment status. This stage will require approximately 300 labelled examples per category from EHRs. The dataset will follow a 60/20/20 schedule including a training (60% of EHRs), validation (20% of EHRs), and an additional test set (20% of EHRs).

###### Stage 3 - Text Generation and Refinement

In the final stage, we will transform the output of our entity extraction into a natural, cohesive CC message. We will begin with a template-based approach, where extracted entities create a basic ‘fill-in-the-blanks’ CC. To further refine this output, we will employ prompt engineering techniques through our generative model, focusing on two key elements: persona and tone. For persona, we will provide the model with specific professional roles to take on (i.e., ‘you are a clinical social worker supporting a patient following a hospitalization). For tone, we will guide the model’s communication style with explicit instructions (i.e., ‘write compassionately while maintaining professional boundaries’). We will also use the CC messages generated in phase 1 of the study as guiding examples for the generative model.

This proposed methodology creates a pipeline that combines entity recognition with language generation, resulting in CC messages that are both professionally appropriate and engaging to the target audience. All model development and testing will be carried out on the high-performance computing cluster managed by Sunnybrook Research Institute. This ensures that no patient information is transferred outside of the Sunnybrook firewall.

Once trained, the LLM will be given the same sample patient EHRs given to staff in phase 1 and will be asked to craft 5 CC messages (1 per EHR). Prior to the commencement of phase 3, study personnel will review all AI and control CC messages to ensure no factual errors are made when generating content from the EHRs. Only identified errors will be changed but no other elements of the CC messages.

**Figure 2.**
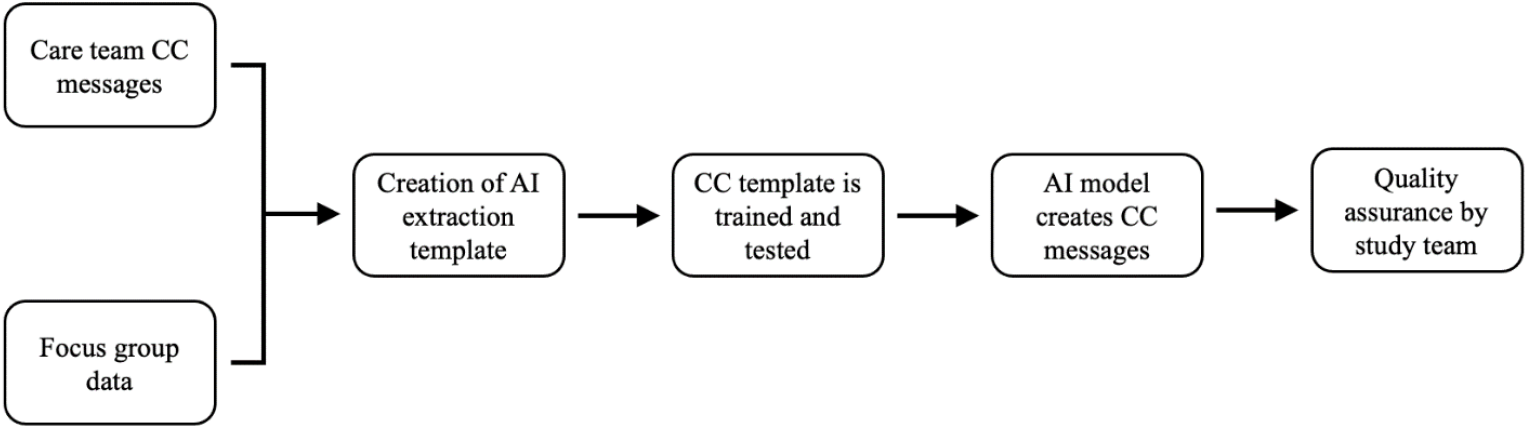
Phase 2 Procedures. *Note*. Procedures to be completed during phase 2 of the study.

#### Phase 3

##### Participants

A sample of 30 PWLE of a psychiatric hospitalization will be recruited to participate. The selection of this study population is to ensure that the perspective of the lived experience will be integrated into any changes made to the CC protocol. Given the potentially sensitive nature of topics discussed in the CC messages, we will ask that participants be at least 1 year since their last discharge from a psychiatric hospitalization.

##### Inclusion Criteria

For inclusion in this phase of the project, participants must be aged 18 or above, able to read and understand English, and have lived experience of psychiatric hospitalization with at least 1 year having passed since last discharge from psychiatric hospitalization.

##### Recruitment

Posters with contact information will be displayed throughout the psychiatry department for potential participants to self-refer. Patients may also be sent information about this study via email through their regular appointment reminders. This study will also be publicized by our Patient and Family Advisory committee (PFAC) as well as Hope and Me, our community partner, via their newsletter and social media platforms.

##### Procedure

Participants will have the option to complete this study online or in-person (i.e., survey will be delivered via REDCap). Following consent procedures, participants will be presented with 10 sample CC messages: 5 created by the LLM and 5 control messages. All information will be de-identified and screened by a study team member to avoid potentially harmful content from being accessed. Participants will be asked to individually rate each CC message on structural characteristics (e.g., style, flow, length) and content of CC message (e.g., tone of message, emotional resonance, relevancy, resources helpful or not). After rating all 10 CC messages, participants will be made aware that AI created some of these messages and will be asked to categorize them based on perceived authorship. Finally, participants will be asked to complete a short questionnaire on the perceptions around the acceptability of introducing AI into psychiatric care.

**Figure 3.**
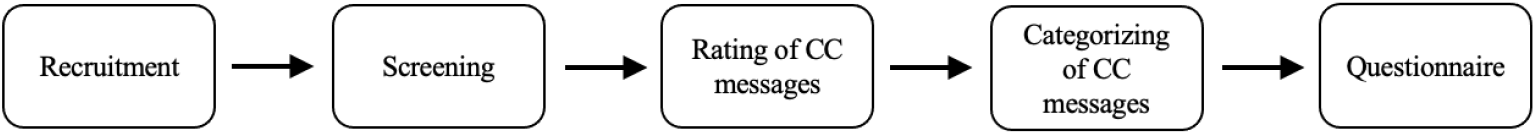
Phase 3 Procedures. *Note*. Procedures to be completed during phase 3 of the study.

### Outcomes and Variables

#### Primary Variables

##### Feedback questionnaire

Feedback questionnaire responses using Likert scales will be quantified and compared in phase 3 to assess acceptability of AI-generated CC messages. This questionnaire will assess the tone, content, and quality of messages made by AI and clinical care staff members.

##### Focus group responses

We will use qualitative thematic analysis to evaluate the focus group transcripts during phase 1 of this project.

#### Demographic Variables

At the time of enrolment for phases 1 and 3, the following demographic information will be obtained: informed consent, full name, age, gender, and sex. For the phase 3 of the project, psychiatric history (i.e., date of last admission, total number of admissions, etc.) will also be obtained.

### Statistical Analysis and Sample Size

For the first phase of the project, we will recruit 10 participants and divide into two focus groups. We will use qualitative thematic analyses for the focus group data. Feedback questionnaire responses will be quantified. The sample size of 10 is in accordance with guidelines for qualitative research and exceeds the necessary population size needed for data saturation.^19^

For the second phase, we will use a sample of approximately 100-200 de-identified patient electronic health records, a small dataset in the context of model development, to train the LLM. This sample is needed to complete name entity recognition, a task that will enable relevant information to be pulled from patient electronic health records for insertion into CC messages. In the process of LLM development, we will measure the correlation between training data sample size and model accuracy to further justify the size of this dataset.

For the final phase we will aim to recruit 30 PWLE participants. The sample size of 30 individuals is in line with recommendations for pilot trials examining interventions with a predicted small to medium effect size.^20^ One-way, within-subjects analyses of variance (ANOVA) will be used to evaluate differences in scores between the two message types on all composite factors. Given the small sample size, we will also use descriptive statistics to compare composite scores of both message types. We will thematically analyse open-ended feedback.

### Outputs and Dissemination Plans

The acceptability of AI and uses of LLMs in the context of mental health care is an area of research that is especially relevant given recent technological advances. This study is one of the first to explore the use of AI for post-discharge messaging in the context of feasibility and acceptability. This research will be helpful for continuing to implement CC messages for inpatient psychiatric uses and other healthcare settings. Personalized CC messaging may help to promote treatment adherence, improve health literacy, and strengthen connections to care teams. Although emerging evidence suggests there are benefits of using AI, such as lowering costs and lessening time constraints, there are ethical concerns that remain. Additional work is needed to determine how to incorporate privacy and safety checks to help monitor the AI-generated content into the clinical workflow in psychiatry settings and beyond.

## Data Availability

All data produced in the present study are available upon reasonable request to the authors, as this is a registered report. Collection of data is ongoing and will be published at a later date.

## Notes

### Competing Interest Statement

All authors have completed the ICMJE uniform disclosure form. The corresponding author declares this project was supported by the Sunnybrook AFP Association through the Innovation Fund of the Alternative Funding Plan from the Academic Health Sciences Centres of Ontario. All authors declared no financial relationships with any organizations that might have an interest in the submitted work in the previous three years; no other relationships or activities that could appear to have influenced the submitted work.

### Funding Statement

This project was supported by the Sunnybrook AFP Association through the Innovation Fund of the Alternative Funding Plan from the Academic Health Sciences Centres of Ontario.

### Author Declarations

The Research Ethics Board of Sunnybrook Health Sciences Centre gave ethical approval for this work.

## References

1. Chung DT, Ryan CJ, Hadzi-Pavlovic D, Singh SP, Stanton C, Large MM. Suicide Rates After Discharge From Psychiatric Facilities. JAMA Psychiatry. 2017;74(7):694–702. doi:10.1001/jamapsychiatry.2017.1044

2. Olfson M, Wall M, Wang S, et al. Short-term Suicide Risk After Psychiatric Hospital Discharge. JAMA Psychiatry. 2016;73(11):1119–1126. doi:10.1001/jamapsychiatry.2016.2035

3. Aaltonen K, Sund R, Hakulinen C, Pirkola S, Isometsä E. Variations in Suicide Risk and Risk Factors After Hospitalization for Depression in Finland, 1996-2017. JAMA Psychiatry. 2024;81(5):506–515. doi:10.1001/jamapsychiatry.2023.5512

4. Adair CE, McDougall GM, Mitton CR, et al. Continuity of Care and Health Outcomes Among Persons With Severe Mental Illness. Psychiatr Serv. 2005;56(9):1061–1069. doi:10.1176/appi.ps.56.9.1061

5. Carter GL, Clover K, Whyte IM, Dawson AH, D’Este C. Postcards from the EDge: 5-year outcomes of a randomised controlled trial for hospital-treated self-poisoning. Br J Psychiatry J Ment Sci. 2013;202(5):372–380. doi:10.1192/bjp.bp.112.112664

6. Comtois KA, Kerbrat AH, DeCou CR, et al. Effect of Augmenting Standard Care for Military Personnel With Brief Caring Text Messages for Suicide Prevention: A Randomized Clinical Trial. JAMA Psychiatry. 2019;76(5):474–483. doi:10.1001/jamapsychiatry.2018.4530

7. Hassanian-Moghaddam H, Sarjami S, Kolahi AA, Lewin T, Carter G. Postcards in Persia: A Twelve to Twenty-four Month Follow-up of a Randomized Controlled Trial for Hospital-Treated Deliberate Self-Poisoning. Arch Suicide Res Off J Int Acad Suicide Res. 2017;21(1):138–154. doi:10.1080/13811118.2015.1004473

8. Skopp NA, Smolenski DJ, Bush NE, et al. Caring contacts for suicide prevention: A systematic review and meta-analysis. Psychol Serv. 2023;20:74–83. doi:10.1037/ser0000645

9. Holman S, Steinberg R, Sinyor M, et al. Caring Contacts to Reduce Psychiatric Morbidity Following Hospitalization During the COVID-19 Pandemic: A Pilot Randomized Controlled Trial. Can J Psychiatry. 2023;68(3):152–162. doi:10.1177/07067437221121111

10. Steinberg R, Amini J, Sinyor M, Mitchell RHB, Schaffer A. Implementation of caring contacts using patient feedback to reduce suicide□related outcomes following psychiatric hospitalization. Suicide Life Threat Behav. Published online June 27, 2024:sltb.13108. doi:10.1111/sltb.13108

11. Milne-Ives M, Selby E, Inkster B, Lam C, Meinert E. Artificial intelligence and machine learning in mobile apps for mental health: A scoping review. PLOS Digit Health. 2022;1(8):e0000079. doi:10.1371/journal.pdig.0000079

12. Domingo J, Galal G, Huang J, et al. Preventing Delayed and Missed Care by Applying Artificial Intelligence to Trigger Radiology Imaging Follow-up. NEJM Catal. 2022;3(4):CAT.21.0469. doi:10.1056/CAT.21.0469

13. Garcia P, Ma SP, Shah S, et al. Artificial Intelligence–Generated Draft Replies to Patient Inbox Messages. JAMA Netw Open. 2024;7(3):e243201. doi:10.1001/jamanetworkopen.2024.3201

14. Murdoch B. Privacy and artificial intelligence: challenges for protecting health information in a new era. BMC Med Ethics. 2021;22(1):122. doi:10.1186/s12910-021-00687-3

15. Boucher EM, Harake NR, Ward HE, et al. Artificially intelligent chatbots in digital mental health interventions: a review. Expert Rev Med Devices. 2021;18(Sup1):37–49. doi:10.1080/17434440.2021.2013200

16. Devlin J, Chang MW, Lee K, Toutanova K. BERT: Pre-training of Deep Bidirectional Transformers for Language Understanding. Published online May 24, 2019. Accessed November 14, 2023. http://arxiv.org/abs/1810.04805

17. Souza F, Nogueira R, Lotufo R. Portuguese Named Entity Recognition using BERT-CRF. Published online February 27, 2020. doi:10.48550/arXiv.1909.10649

18. Touvron H, Martin L, Stone K, et al. Llama 2: Open Foundation and Fine-Tuned Chat Models. Published online July 19, 2023. doi:10.48550/arXiv.2307.09288

19. Hennink M, Kaiser BN. Sample sizes for saturation in qualitative research: A systematic review of empirical tests. Soc Sci Med 1982. 2022;292:114523. doi:10.1016/j.socscimed.2021.114523

20. Whitehead AL, Julious SA, Cooper CL, Campbell MJ. Estimating the sample size for a pilot randomised trial to minimise the overall trial sample size for the external pilot and main trial for a continuous outcome variable. Stat Methods Med Res. 2016;25(3):1057–1073. doi:10.1177/0962280215588241

21. Graham, S., Depp, C., Lee, E. E., Nebeker, C., Tu, X., Kim, H.-C., & Jeste, D. V. (2019). Artificial Intelligence for Mental Health and Mental Illnesses: an Overview. Current Psychiatry Reports, 21(11), 116–118. 10.1007/s11920-019-1094-0

22. Alammar, J., & Grootendorst, M. (2024). Hands-on large language models: language understanding and generation (First edition.). O’Reilly Media, Inc.

